# A Recursive Bifurcation Model for Predicting the Peak of COVID-19 Virus Spread in United States and Germany

**DOI:** 10.1101/2020.04.09.20059329

**Authors:** Julia Shen

## Abstract

Prediction on the peak time of COVID-19 virus spread is crucial to decision making on lockdown or closure of cities and states. In this paper we design a recursive bifurcation model for analyzing COVID-19 virus spread in different countries. The bifurcation facilitates a recursive processing of infected population through linear least-squares fitting. In addition, a nonlinear least-squares fitting is utilized to predict the future values of infected populations. Numerical results on the data from three countries (South Korea, United States and Germany) indicate the effectiveness of our approach.

## 1. Introduction

Coronavirus disease (COVID-19) is a novel respiratory illness that originated in 2019 and can spread from person to person, as defined by CDC [1]. The first incidence of such disease was publicly reported as an outbreak in Wuhan, China. So far, the original source of this disease has not been clearly identified and the disease is continuously spread in over 70 countries.

Lockdown of towns, cities, states, and countries imposes severe damage to the well-being and economic growth of society. The unknown nature about the peak of virus spread makes the decision of lockdown or closure a difficult task to plan in advance. This calls for an accurate model to predict the peak time of ongoing spread of COVID-19 virus.

## 2. Literature Review

Many studies have been carried out on the epidemic investigation of COVID-19 spread. The first category of studies is pure statistical analysis. Important epidemic parameters were estimated [2, 3], including basic reproduction number [4], doubling time [5] and serial interval [6]. In addition, some advanced models were developed in handling untraced contacts [7], undetected international cases [8], and actual infected cases [9]. Statistical reasoning [10, 11] and stochastic simulation [12, 13] were also explored by a few researchers.

The second group of investigations was based on dynamic modelling. Susceptible exposed infectious recovered model (SEIR) was used in assessing various measures in the COVID-19 outbreak [14-17]. Furthermore, it was utilized in investigating the effect of lockdown [18], transmission process [19], transmission risk [14], and the effect of quarantine [14]. The SEIR model with time delays was also developed for studying the period of incubation and recovery [20, 21].

Although there have been many recent studies with respect to the COVID-19 virus spread, an accurate model to pinpoint the peak time of the virus spread is still elusive. Such a model is crucial to a decision-making process for strategic plans to achieve a balance between reduction in life loss and avoidance of economic crisis due to lockdown.

The rest of this paper is organized as follows. In Section 3, a recurve bifurcation model is introduced to model the COVID-19 spread. A bifurcation analysis is given in Section 4 on infected data from South Korea. Section 5 describes the prediction of COVID-19 virus based on our model, followed by some concluding remarks in Section 6.

## 3. Recursive Bifurcation Model

In this paper, we focus on the number of infected population, which is an important metric to measure the extent of the COVID-19 spread in different countries. Although the infected population in most countries follows a pattern of an exponential or sigmoid function, the logarithm of the infected population may provide more information, as shown in Fig. 1(b).

**Figure 1:**
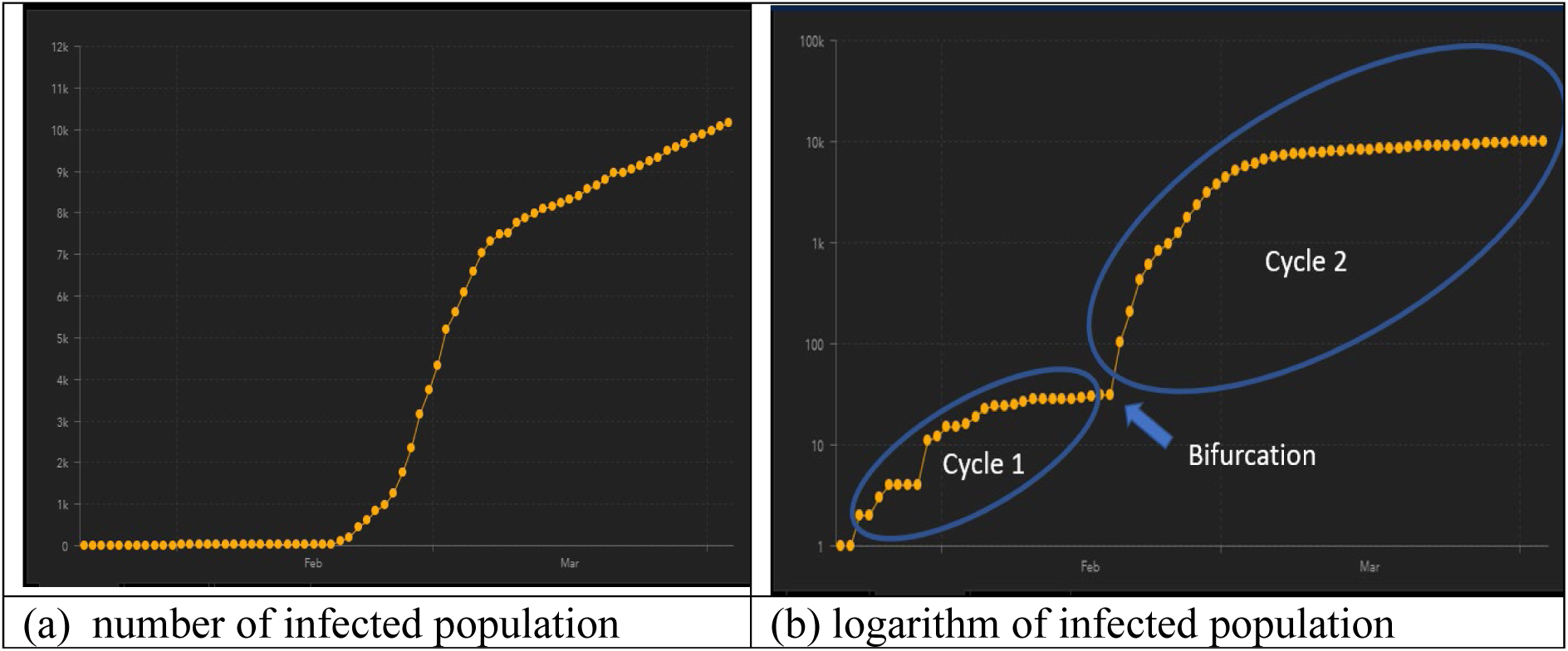
The number of infected population in South Korea as of April 5, 2020.

The countries that exhibit a bifurcation pattern include South Korea, United States, France, Canada, Germany, Australia, Malaysia, and Ecuador. By utilizing the bifurcation, we can find out the intrinsic parameters in cycle 1 and apply those parameters as a set of starting values in the prediction for cycle 2 or beyond.

Following the above idea, we introduce a recursive Tanh function to describe the number of infected population within each cycle of an entire virus spread process:

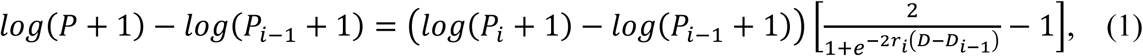

where *i* refers to the *i*-th cycle, *P* is the number of infected population in the *i*-th cycle, *D* represents the number of days since the initiation of virus spread, *P*_*i*_ stands for the number of infected population at the end of the *i*-th cycle, *r*_*i*_ is the spread rate in the *i*-th cycle, and *D*_*i*_ refers to the number of days at the end of the *i*-th cycle. The purpose of adding 1 in the logarithm calculation is to avoid an infinity caused by the case where *P* = 0.

Note that Equation (1) is not strictly a recursive formula in a conventional sense. The reason for us to call it as a recursive one is that Equation (1) should be recursively solved starting from cycle 1 toward cycle *n*, if *n* is the last cycle for the virus spread. When *n*=1, this equation is degenerated to a regular Tanh function.

## 4. Bifurcation Analysis of COVID-19 Virus Spread

In order to validate Equation (1) for the analysis of COVID-19 virus spread, we have to select a complete virus spread process. Among all the countries, South Korea seems to be the best choice for this validation because the country provides reasonably reliable data and the virus spread in that country has been stabilized.

*r*_*i*_ in Equation (1) represents an intrinsic attribute of the virus spread rate. It can be estimated by a linear least-squares fitting of the following linear equation in a parameter space:

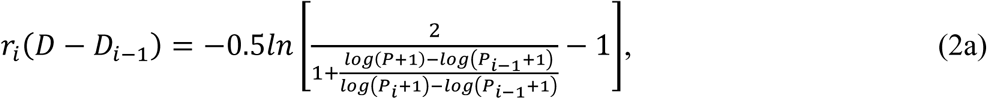

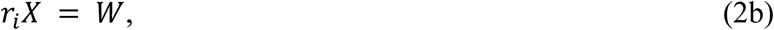

where *X* = *D*-*D*_*i*_ and 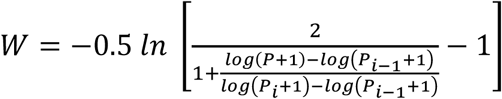.

Figure 2(a) shows the result of determining the virus spread rate, *r*_1_. By using this *r* value, we predict the infected population, *y*_*p*_, which is very close to the true data, *y*, as shown in Figure 2(b).

**Figure 2:**
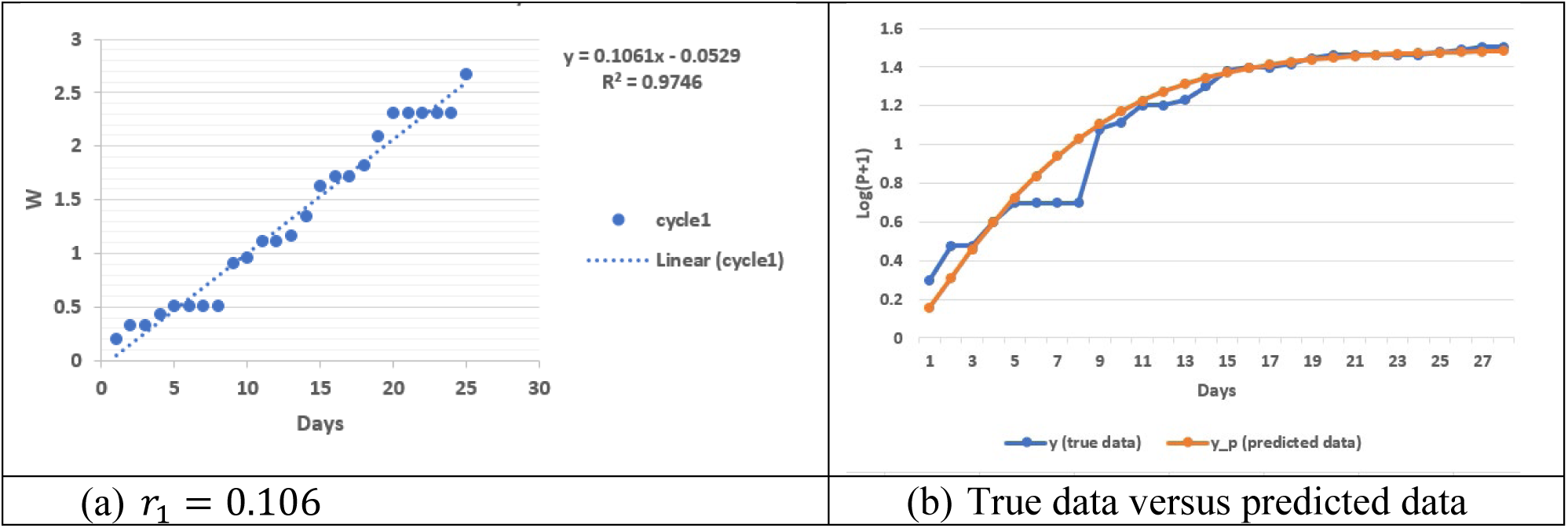
Determination of virus spread rate with South Korea data in cycle 1.

Furthermore, by using *r*_1_ in cycle 2 of Korea data, we also achieve an accurate prediction of infected population and validate *α* to be close to unity (Figure 3). Here, *α* is a fictious variable that should be of a value of unity:

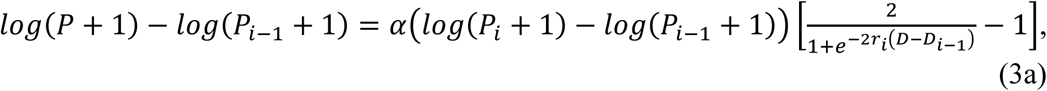

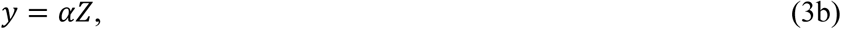

where

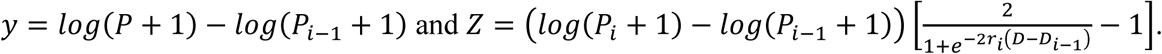

The bifurcation in Figure 1(a) is easy to identify visually. An automatic algorithm can be created on the basis of discontinuity of tangential direction when traversing the curve. Since it is not the main focus of this paper, we do not explore it any further in this aspect.

**Figure 3:**
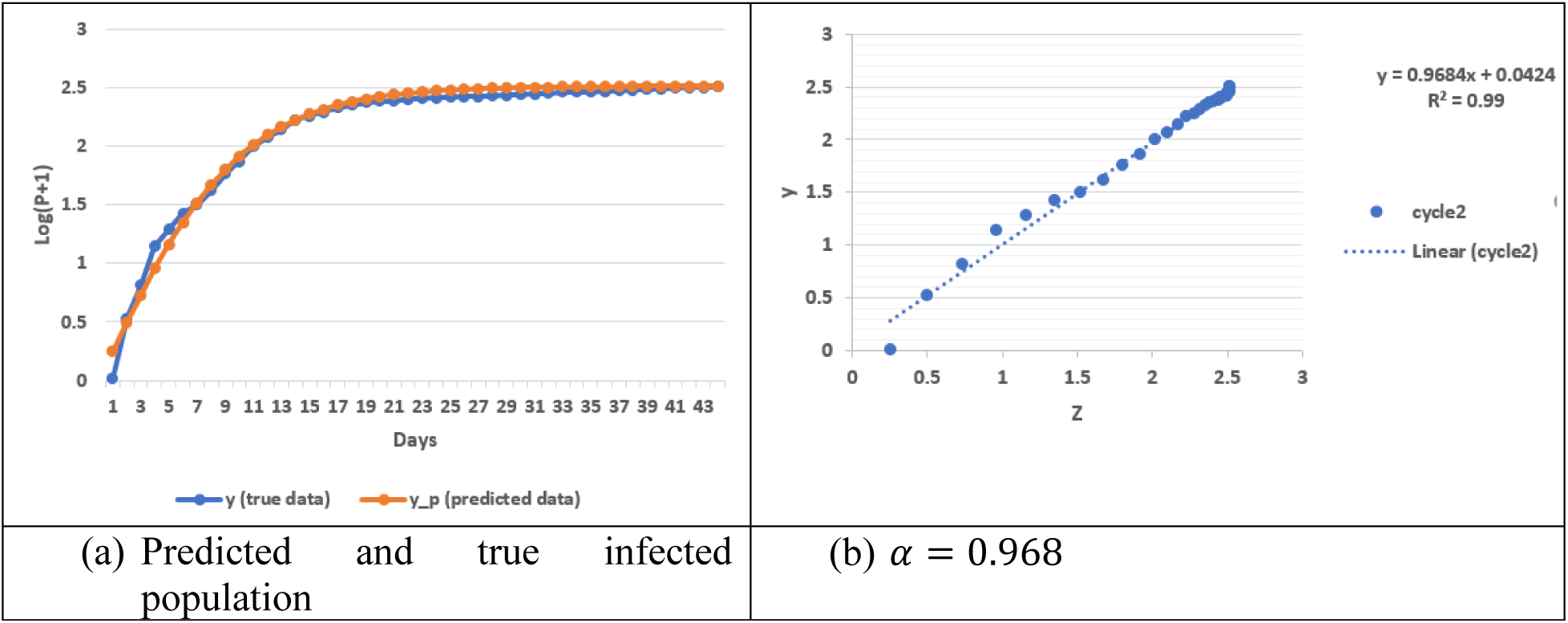
Analysis of virus spread with South Korea data in cycle 2.

## 5. Prediction of Peak Time of COVID-19 Virus Spread

Based on the model in Section 4, we design an algorithm to predict the incoming peak time of COVID-19 virus in United States and Germany, as given in Table 1. Since the infected population has not been stabilized in these two countries, it is important to estimate the ultimate infected population at the end of the last cycle, *n*.

**Table 1:** An Algorithm for Predicting the Peak Time of COVID-19 Virus Spread

|  |  |
| --- | --- |
| Step 1 | Determine the virus spread rate in cycle 1, $r_1$, based on a least-squares fitting of Equation (2) |
| Step 2 | Recursively analyze the infected population in cycles 2 through $n-1$. |
| Step 3 | Estimate the virus spread rate in cycle $n$, $\hat{r}_n$, based on a linear least-squares fitting of Equation (2) |
| Step 4 | Estimate the logarithm of infected population in cycle $n$ by a linear least-squares fitting of $\hat{\beta}_n$ in Equation (4) |
| Step 5 | Determine $\beta_n$ and $r_n$ by using a nonlinear Levenberg-Marquart least-squares fitting based on $[\hat{r}_n, \hat{\beta}_n]$ as a pair of starting values through Equation (5) |
| Step 6 | Predict the future infected population based on Equation (5) |
| Step 7 | Use a termination condition (Eq. 6) to estimate the peak time of virus spread |

We first use the following formula to estimate 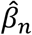 through a linear least-squares fitting:

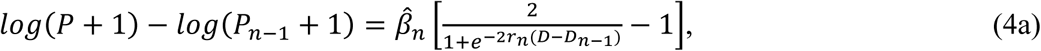

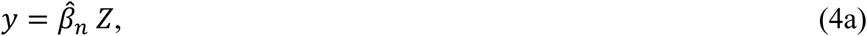

where *y*=*log*(*P*+1)-*log*(*P*_*n*-1_+1) and 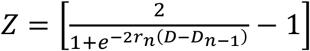.

Then, Equation (1) is utilized to estimate 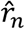 for cycle *n* through a linear least-squares fitting. With 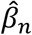 and 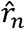 being available as a pair of starting values, a nonlinear Levenberg-Marquart least-squares fitting [22] is computed to determine two unknown parameters (*β*_*n*_ and *r*_*n*_) simultaneously in the following equation:

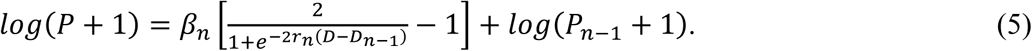

Once *β*_*n*_ and *r*_*n*_ are determined, Equation (5) can be used to predict the future values of infected population. To define the peak time of virus spread, a termination condition is proposed as follows:

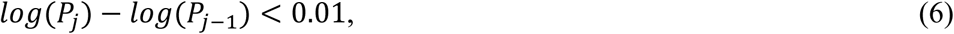

where *j* refers to *j*-th day in cycle *n*. Equation (6) means that the virus spread approaches its stability when the difference in the logarithm of infected population between two consecutive days is less than 0.01.

Figure 4 shows the prediction result of infected population in United States. The bifurcation pattern of infected population is given in Figure 4(a) and the determination of virus spread rate is presented in Figure 4(b). The virus spread rate in United States (*r*_1_ = 0.072) is smaller than that in South Korea (*r*_1_ = 0.106) because the population density in South Korea is much higher. This may also mean that the peak time of virus spread will be longer than that in South Korea. Figures 4(c) and 4(d) are the predicted data for cycles 1 and 2, respectively. According to Figure 4(d), the COVID virus spread in United States will roughly peak on April 26, 2020.

**Figure 4:**
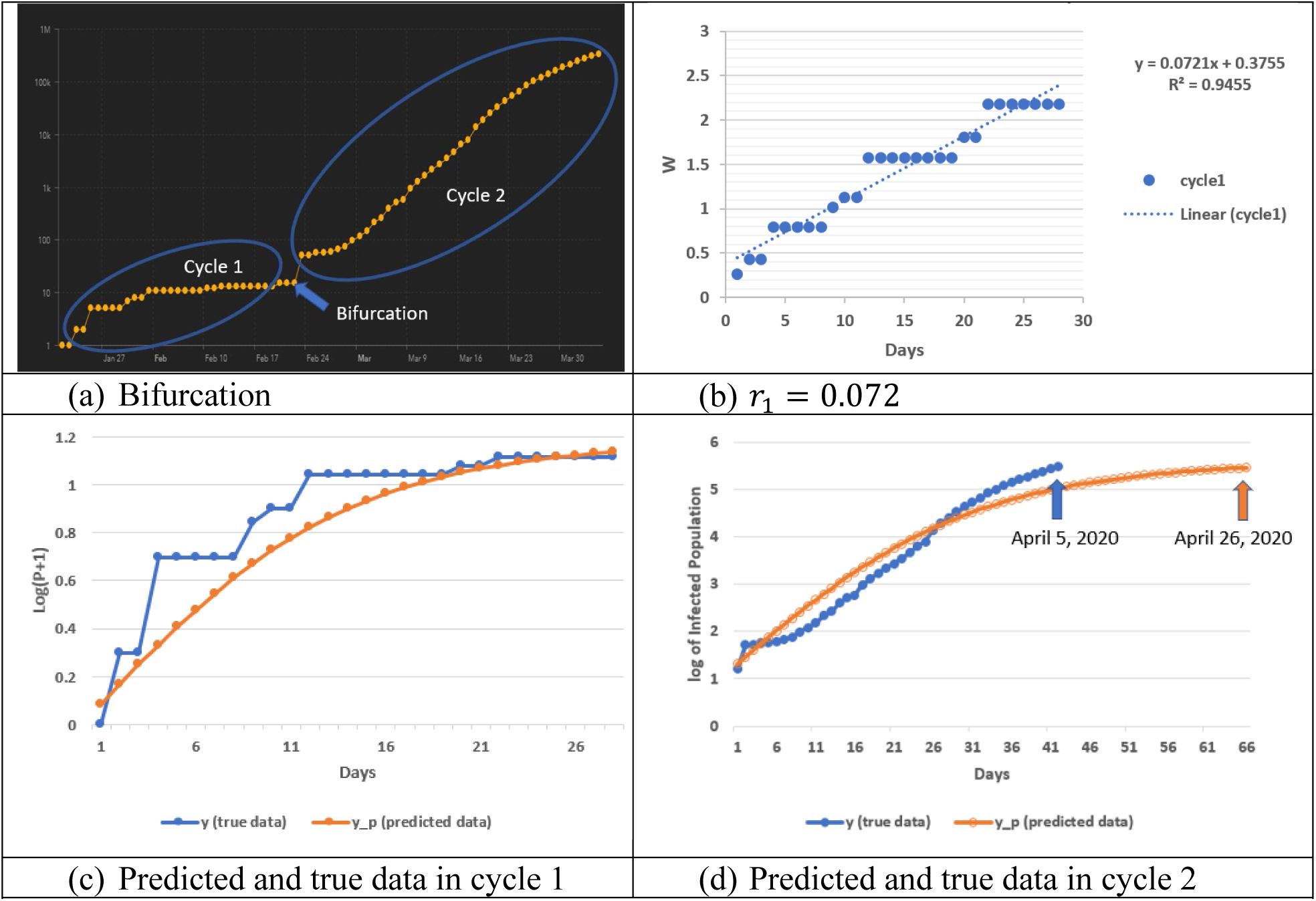
Prediction of the peak time of COVID-19 spread in United States.

COVID-19 data in Germany can be analyzed in a similar way. Figure 5(b) indicates that the virus spread will approximately peak on May 1, 2020. The virus spread rate, *r*_1_, in Germany is 0.108, which is close to that in South Korea. These two countries have a higher virus spread rate than United State because of the higher population density in Germany and South Korea.

**Figure 5:**
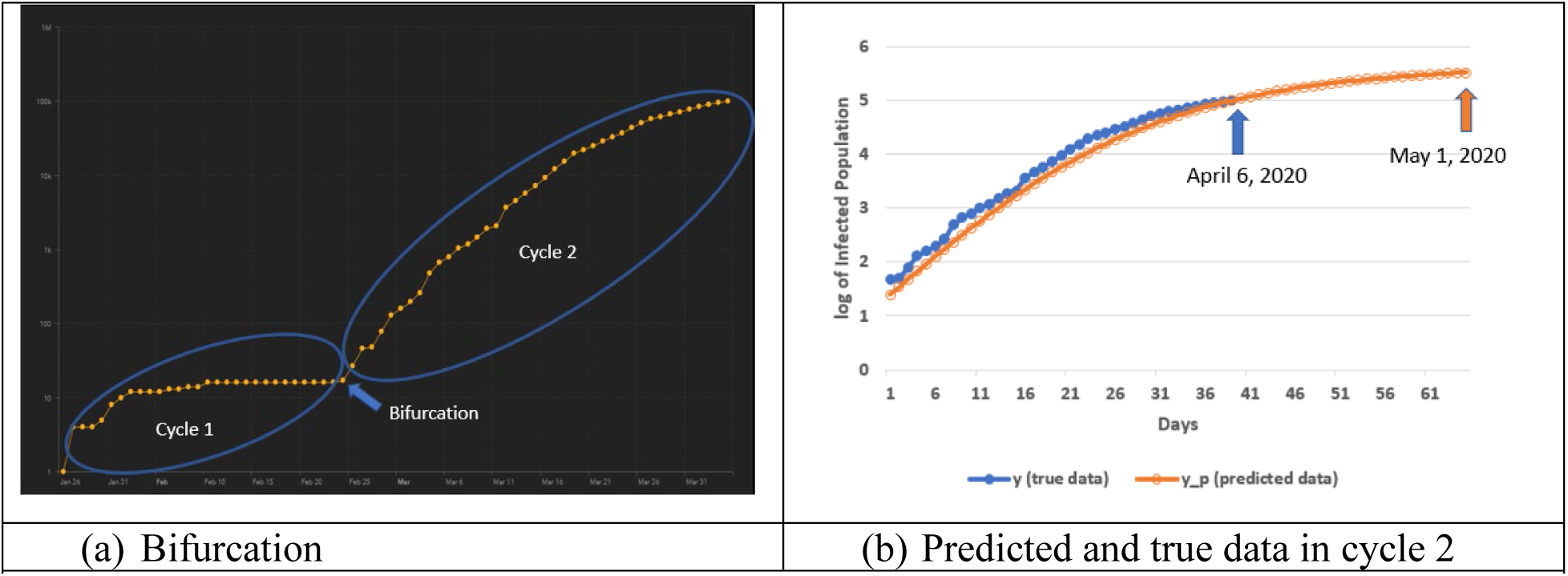
Prediction of the peak time of COVID-19 spread in Germany.

## 6. Conclusions

In this paper, we propose a recursive bifurcation approach to estimate the peak time of COVID-19 virus spread. The infected population data in South Korea is analyzed as an example of stabilized virus spread. An algorithm is developed to predict the future infected population based on ongoing existing data as of April 6, 2020. Our model predicts that the COVID-19 virus spread will approximately peak on April 26 and May 1, 2020, respectively for United States and Germany in terms of infected population.

## Data Availability

All the true data of infected populations was obtained from the Coronavirus Resource Center of Johns Hopkins University.

https://coronavirus.jhu.edu/map.html

## Acknowledgments

All the true data of infected populations is obtained from the Coronavirus Resource Center of Johns Hopkins University.

## References

[1] CDC. What you need to know about coronavirus disease 2019 (COVID-19). Centers for Disease Control and Prevention (cdc.gov/COVID19), Atlanta, Geogia, U.S.A. 2020.

[2] Sanche S, Lin YT, Xu C, Romero-Severson E, Hengartner N, Ke R. The novel coronavirus 2019-ncov is highly contagious and more infectious than initially estimated. medRxiv. 2020; (2002.03268).

[3] Lai S, Bogoch I, Ruktanonchai N, Watts A, Li Y, Yu J, Lv X, Yang W, Yu H, Khan K, Li Z, Tatem AJ. Assessing spread risk of Wuhan novel coronavirus within and beyond China, January-April 2020: a travel network-based modelling study. medRxiv. 2020; (doi:10.1101/2020.02.04.20020479).

[4] Zhao S, Lin Q, Ran J, Musa SS, Yang G, Wang W, Lou Y, Gao D, Yang L, He D, Wang MH. Preliminary estimation of the basic reproduction number of novel coronavirus (2019-ncov) in China, from 2019 to 2020: A data-driven analysis in the early phase of the outbreak. International Journal of Infectious Diseases. 2020; 92:214–217.

[5] Muniz-Rodriguez K, Chowell G, Cheung CH, Jia D, Lai PY, Lee Y, Liu M, Ofori SK, Roosa KM, Simonsen L, Fung IC. Epidemic doubling time of the 2019 novel coronavirus outbreak by province in mainland China. medRxiv. 2020; (http://dx.doi.org/10.1101/2020.02.05.20020750).

[6] Nishiura H, Linton NM, Akhmetzhanov AR. Serial interval of novel coronavirus (2019-ncov) infections. International Journal of Infectious Diseases. 2020; 93(2020):284–286.

[7] Nishiura H, Jung S, Linton NM, Kinoshita R, Yang Y, Hayashi K, Kobayashi T, Yuan B, Akhmetzhanov AR. The extent of transmission of novel coronavirus in Wuhan, China, 2020. Journal of Clinical Medicine. 2020; 9(2):330-1-330-5.

[8] De Salazar PM, Niehus R, Taylor A, Buckee CO, Lipsitch M. Using predicted imports of 2019-ncov cases to determine locations that may not be identifying all imported cases. medRxiv. 2020; (doi: https://doi.org/10.1101/2020.02.04.20020495).

[9] Zhao H, Man S, Wang B, Ning Y. Epidemic size of novel coronavirus-infected pneumonia in the epicenter Wuhan: using data of five-countries’ evacuation action. medRxiv. 2020; (doi: https://doi.org/10.1101/2020.02.12.20022285).

[10] Chinazzi M, Davis JT, Ajelli M, Gioannini C, Litvinova M, Merler S, Piontti AP, Rossi L, Sun K, Viboud C, Xiong X, Yu H, Halloran ME, Longini IM, Vespignani A. The effect of travel restrictions on the spread of the 2019 novel coronavirus (2019-ncov) outbreak. medRxiv. 2020; (doi: https://doi.org/10.1101/2020.02.09.20021261).

[11] Jin C, Yu J, Han L, Duan S. The impact of traffic isolation in Wuhan on the spread of 2019-nov.medRxiv. 2020; (doi: https://doi.org/10.1101/2020.02.04.20020438).

[12] Hellewell J, Abbott S, Gimma A, Bosse N, Jarvis CI, Russel TW, Munday JD, Kucharski AJ, Edmunda WJ, CMMID nCoV working group, Funk S, Eggo RM. Feasibility of controlling 2019-ncov outbreaks by isolation of cases and contacts. THE LANCET Global Health. 2020; 8(4):e488–e496.

[13] Quilty B, Clifford S, Flasche S, Eggo RM. Effectiveness of airport screening at detecting travellers infected with 2019-ncov. Eurosurveillance. 2020; 25(5):1–6.

[14] Tang B, Bragazzi NL, Li Q, Tang S, Xiao Y, Wu J. An updated estimation of the risk of transmission of the novel coronavirus (2019-ncov). Infectious Disease Modelling. 2020; 5(2020):248–255.

[15] Shen M, Peng Z, Guo Y, Xiao Y, Zhang L. Lockdown may partially halt the spread of 2019 novel coronavirus in Hubei province, China. medRxiv. 2020; (doi: https://doi.org/10.1101/2020.02.11.20022236).

[16] Clifford SJ, Klepac P, Zandvoort KV, Quilty BJ, Eggert D, Flasche S. Interventions targeting air travellers early in the pandemic may delay local outbreaks of sars-cov-2. medRxiv. 2020; (doi: https://doi.org/10.1101/2020.02.12.20022426.).

[17] Xiong H, Yan H. Simulating the infected population and spread trend of 2019-ncov under different policy by EIR model. medRxiv. 2020; (https://doi.org/10.1101/2020.02.10.20021519).

[18] Li X, Zhao X, Sun Y. The lockdown of Hubei province causing different transmission dynamics of the novel coronavirus (2019-ncov) in Wuhan and Beijing. medRxiv. 2020; (doi: https://doi.org/10.1101/2020.02.09.20021477).

[19] Chen T, Rui J, Wang Q, Zhao Z, Cui JA, Yin L. A mathematical model for simulating the transmission of Wuhan novel coronavirus. bioRxiv. 2020; (doi: https://doi.org/10.1101/2020.01.19.911669).

[20] Yue Y, Chen Y, Liu K, Luo X, Xu B, Jiang Y, Cheng J. Modeling and prediction for the trend of outbreak of NCP based on a time-delay dynamic system. Scientia Sinica Mathematica. 2020; 50(3):1–8.

[21] Chen Y, Cheng J, Jiang Y, Liu K. A time delay dynamical model for outbreak of 2019-ncov and the parameter identification. medRxiv. 2020; (2002.00418).

[22] Press WH, Teukolsky SA, Vetterling WT, Flannery BP. Numerical Recipes in C: The Art of Scientific Computing. Cambridge University Press: Cambridge,1992; 994 pp.

